# Moderate-to-vigorous physical activity, age-dependent adiposity, and obesity risk in older adults at high cardiovascular risk

**DOI:** 10.64898/2026.09.14.26362982

**Authors:** Eleonora Fornara, Marta H. Hernández, Alberto Goday, Anna Camps-Vilaró, Miguel Ángel Martínez-González, Jordi Salas-Salvadó, Ramón Estruch, Emilio Ros, Raquel Cueto-Galán, Miquel Fiol, José Lapetra, Juan Timiraos, Lluís Serra-Majem, Xavier Pintó, Zenaida Vázquez-Ruiz, Francisco Manuel Martín-Luján, Montserrat Fitó, Álvaro Hernáez

## Abstract

**Objective:** To investigate the association between usual moderate-to-vigorous leisure-time physical activity (MVLTPA) and longitudinal trajectories of BMI and waist-to-height ratio (WHtR), as well as the incidence of obesity (BMI ≥30 kg/m^2^) and central adiposity (WHtR ≥0.6), in older adults at high cardiovascular risk.

**Methods:** In PREDIMED participants aged 60-80 years (61% women; n=5,273 for BMI, n=4,940 for WHtR), MVLTPA was assessed using the Minnesota questionnaire (MET·min/day, cumulative mean). Mixed-effects models with cubic splines were used to estimate BMI and WHtR trajectories according to MVLTPA levels (≥100 [adequate] vs <100 [insufficient]). Incident obesity outcomes were assessed with Cox models.

**Results:** Participants reporting ≥100 MET·min/day showed lower BMI and WHtR values compared with less active individuals (differences ranging from-1.27 to-1.48 kg/m^2^ and-0.021 to-0.026, respectively), and slower declines in BMI. Adequate MVLTPA was associated with a 34% lower risk of central adiposity (HR 0.66 [0.56; 0.77]) and a 24% lower risk of obesity only in women (HR 0.76 [0.57; 1.00], *p*-interaction = 0.084). Nonlinear analyses suggested that central adiposity risk decreased up to 400 MET·min/day and plateaued thereafter.

**Conclusions:** Higher MVLTPA is associated with favourable adiposity trajectories and lower risk of central adiposity in older adults at high cardiovascular risk.

**Study Importance:** *What is already known?:*

- Obesity, particularly central adiposity, is a major and modifiable cardiovascular risk factor, and regular physical activity is widely recommended as a cornerstone for obesity prevention.
- Evidence in older adults remains limited, particularly regarding long-term adiposity trajectories, as most available data come from short-term randomized controlled trials focused on changes in BMI and waist circumference. There is little evidence from longer follow-up studies examining complementary biomarkers such as waist-to-height ratio (WHtR), sex-specific associations, clinically relevant incident obesity-related outcomes, or the dose of moderate-to-vigorous leisure-time physical activity (MVLTPA) associated with the lowest adiposity risk.

*What does this study add?:*

- In older adults at high cardiovascular risk, MVLTPA ≥100 MET·min/day was associated with lower predicted BMI and WHtR values between 60 and 80 years of age, slower late-life BMI decline, and a 34% lower risk of incident central adiposity.
- Dose-response analyses showed a significant, nonlinear inverse association between MVLTPA and central adiposity risk, with progressively lower risk up to approximately 400 MET·min/day and a plateau thereafter.

*How might your results change the direction of research or the focus of clinical practice?:*

- These findings support sustained MVLTPA as part of cardiovascular prevention in older adults at high risk and highlight WHtR as a useful waist-based marker for monitoring adiposity-related benefits of physical activity.
- The results support promoting achievable increases in physical activity, particularly among inactive or insufficiently active older adults and women, while suggesting that higher MVLTPA volumes may be relevant for central adiposity.

## INTRODUCTION

Cardiovascular diseases remain the leading cause of mortality worldwide (1). Among established cardiovascular risk factors, excess adiposity has a key role, with both overall adiposity, commonly assessed using body mass index (BMI), and central adiposity, captured by indices such as the waist-to-height ratio (WHtR), being associated with adverse cardiometabolic outcomes and mortality (2, 3). In this context, moderate-to-vigorous leisure-time physical activity (MVLTPA) has been associated with a lower weight gain in prospective cohort studies (4), and short-term randomized controlled trials have shown beneficial effects of physical activity on BMI and WHtR (5). To improve these and other risk factors, current guidelines recommend at least 150 minutes/week of moderate-intensity or 75 minutes/week of vigorous-intensity physical activity for adults and older adults to reduce cardiovascular mortality (6, 7), which corresponds to approximately 100 metabolic equivalents of task-minute per day (MET·min/day) of MVLTPA. However, these recommendations are largely uniform across age groups and sexes (8). This is particularly relevant in older adults, in whom the clinical impact of adiposity may be further amplified by age-related changes in body composition, including progressive loss of lean mass due to sarcopenia and visceral fat accumulation (9–11). Evidence from longer follow-up studies examining adiposity and the incidence of clinically relevant obesity-related outcomes is scarce in older adults (12–15). Consequently, important gaps remain, including: 1) the lack of studies describing age-dependent trajectories in this age segment; 2) sex-specific trajectories for women and men; 3) associations among individuals with elevated adiposity; and 4) the precise MVLTPA dose associated with the lowest risk of adverse adiposity outcomes. Addressing these gaps could help refine current physical activity recommendations. Therefore, this study aimed to investigate the association between MVLTPA and the age-dependent evolution of adiposity in older adults at high cardiovascular risk by examining age-related trajectories of BMI and WHtR between ages 60-80, the incidence of overall obesity and central adiposity, and potential dose-response and sex-specific patterns of association.

## METHODS

### Study population

For this work, data from PREDIMED study were used as a prospective cohort. The PREDIMED study was a multicenter, randomized, controlled intervention trial conducted in Spain between 2003 and 2010, aimed at assessing the long-term effects of the Mediterranean diet on the primary prevention of major cardiovascular events in older adults (16). Eligible participants were men aged 55-80 years and women aged 60-80 years, free of cardiovascular disease at enrolment but presenting type 2 diabetes or at least three major cardiovascular risk factors, including smoking, hypertension, dyslipidaemia, overweight or obesity, and family history of premature coronary heart disease. For the present analyses, the study population was restricted to participants aged 60-80 at baseline. As shown in **Figure 1**, participants were excluded from the analyses if they were younger than 60 years at baseline, had missing baseline data on MVLTPA, or anthropometric measurements available only at baseline. For survival analyses, only participants presenting the corresponding condition at baseline were excluded. This study was conducted and reported in accordance with the Strengthening the Reporting of Observational Studies in Epidemiology (STROBE) guidelines.

**Figure 1.**
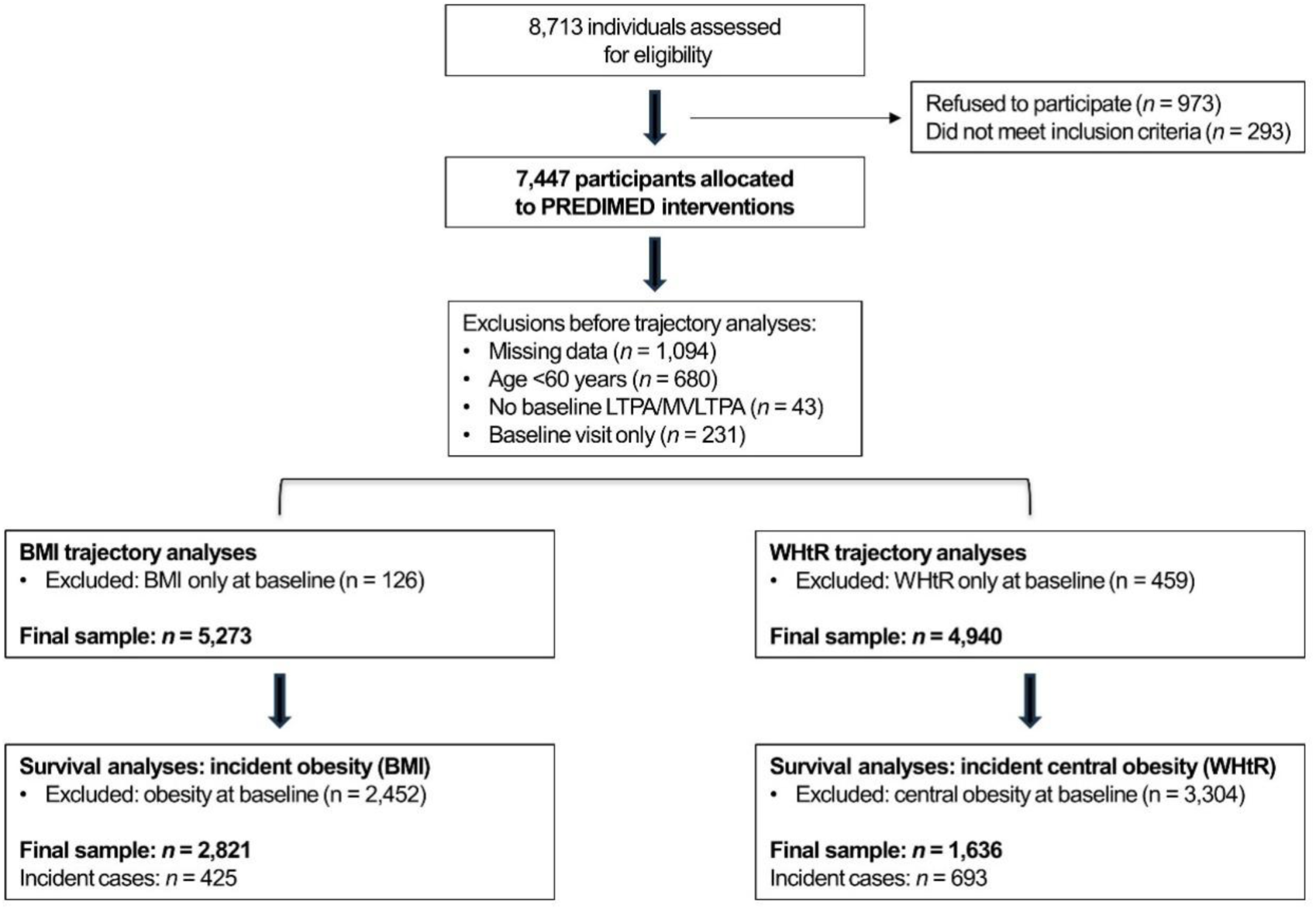
Flow chart

### Exposure: usual moderate-to-vigorous leisure-time physical activity levels

MVLTPA was estimated using the Minnesota Leisure Time Physical Activity Questionnaire, validated in both Spanish men and women (17, 18). Participants reported the frequency (days) and duration (minutes per day) of 67 activities performed during the previous year. MVLTPA was quantified using metabolic equivalents of task-minutes per day (MET·min/day) and calculated by multiplying the MET value assigned to each activity of moderate and vigorous intensity by its average daily duration and summing across all reported activities. MVLTPA was assessed at each study visit, and all available measurements per participant were used to calculate the participant-specific cumulative average exposure. This cumulative average was additionally categorized as adequate (≥100 MET·min/day) or insufficient (<100 MET·min/day, reference) MVLTPA (7).

### Outcomes

Information on anthropometric parameters (weight, waist circumference, and height) was collected by trained clinical staff at baseline and at annual visits between 2003 and 2010. BMI in kg/m^2^ was calculated for each visit using weight and height values, and WHtR (without units) by dividing waist circumference in cm by height in cm. With these data, we constructed an age-indexed dataset in which each BMI and WHtR measurement was assigned to the participant’s chronological age at the time of assessment (12). Thus, participants contributed data only for the specific ages at which they were observed during follow-up. Measurements from different participants, observed at overlapping ages between 60 and 80 years, were then combined to estimate age-related trajectories of BMI and WHtR (12). This approach does not imply that any individual participant was continuously observed throughout the entire age interval. The number of BMI and WHtR observations available for each age is reported in **Supplementary Table 1**.

Based on these measurements, we defined the onset of two clinically relevant conditions: overall obesity (BMI ≥30 kg/m²) and central adiposity (WHtR ≥0.6) (19). Onset was defined as the first occurrence of the condition in participants without the condition at baseline (20).

### Other variables

In the baseline PREDIMED visit, trained clinical staff collected information on age (years), self-reported sex (women/men), recruitment center, educational level (primary, secondary, higher), smoking status (current, former, or never smoker), energy intake (kcal/day, from a validated 137-item food frequency questionnaire), adherence to a Mediterranean diet (using the 14-point Mediterranean Diet adherence short screener by Schroder H et al.) (21), and prevalence of diabetes (yes/no), hypertension (yes/no), and hypercholesterolemia (yes/no). Before the analyses, covariates with non-structural missingness at baseline were imputed using additive regression, bootstrapping, and predictive mean matching based on all available information from the other study variables (1.8% for educational level, 1.0% for energy intake, and 0.7% for adherence to the Mediterranean diet).

### Ethical aspects

This study was conducted in accordance with the principles of the Declaration of Helsinki. The protocol was approved by the Ethics Committee of Parc de Salut Mar (reference 2024/11670/I; November 18, 2024). All participants provided written informed consent before enrolment in the PREDIMED study.

### Statistical analyses

Characteristics of the participants in the baseline PREDIMED visit were summarized using means and standard deviations for normally distributed continuous variables, medians and interquartile ranges for non-normally distributed variables, and proportions for categorical variables.

### Longitudinal trajectory analyses

Age-dependent trajectories, defined as model-predicted mean values of BMI and WHtR at each age, were modelled in the longitudinal aggregated database spanning ages 60-80 years using mixed-effects regression models with multivariable smoothed cubic splines (*K*+4 degrees of freedom) (12, 22, 23). Models included an interaction term between age and MVLTPA category to allow trajectories to differ across groups. Models were adjusted for sex, educational level, PREDIMED intervention group, diabetes, hypertension, hypercholesterolemia, smoking status, adherence to a Mediterranean diet, and energy intake. We graphically represented the mean age-related trajectories of BMI and WHtR according to MVLTPA category using predicted values, and estimated between-group differences at ages 60, 65, 70, 75, and 80 years using linear regression models. BMI and WHtR trajectories were also estimated separately in women and men (after assessing whether trajectories differed by sex using likelihood ratio tests for interaction), and in participants with excess adiposity at baseline (obesity for BMI analyses and central adiposity for WHtR analyses). We additionally estimated the BMI change between ages 70 and 80 years as the slope of the linear equation fitted to the predicted BMI values across this age range.

### Incident outcome analyses

The association between MVLTPA and the risk of developing overall obesity and central adiposity was examined using two complementary approaches. First, we presented the cumulative incidence of each outcome between ages 60 and 80 years across MVLTPA categories with 95% confidence intervals using Kaplan-Meier curves. Age was used as the underlying time scale, with entry at baseline age and exit at the age of event occurrence or the age on December 1, 2010, whichever occurred first. Kaplan-Meier curves were generated in the full study population and separately for women and men (after assessing whether survival models differed by sex using likelihood ratio tests for interaction). To minimize confounding, inverse probability weights were applied; these weights were derived from a propensity score model adjusted for the same set of covariates included in the trajectory analyses. Second, associations between the cumulative average of MVLTPA and the risk of developing obesity or central adiposity were evaluated using age-as-time-scale multivariable Cox proportional hazards models. The models were adjusted for the same covariates as those included in the trajectory analyses. In the first set of analyses, we estimated hazard ratios (HRs) for adequate MVLTPA (≥100 MET·min/day, according to the cumulative average) versus insufficient MVLTPA (<100 MET·min/day, reference). In the second set, we examined dose-response associations with MVLTPA entered as a continuous variable (participants reporting zero MET·min/day were used as the reference group). In these analyses, linearity was assessed by comparing a model including only the linear MVLTPA term with a model additionally including smoothed cubic spline terms, using likelihood ratio tests. When evidence of non-linearity was detected, spline-based Cox models were used to characterize the dose-response association and to identify the MVLTPA value associated with the lowest risk of each obesity outcome; segment-specific estimates were reported for ranges showing the strongest risk gradient. For graphical purposes, MVLTPA values were truncated at 500 MET·min/day. After assessing whether nonlinear models differed by sex using likelihood ratio tests for interaction, we also presented them separately for women and men.

All analyses were conducted using R software (version 4.3.1). Code for data management and statistical analyses is available at https://github.com/alvarohernaez/MVLTPA_cardiometab_PREDIMED

## RESULTS

### Study participants

The study population consisted predominantly of older adults with a high burden of cardiovascular risk factors. The BMI analyses included 5,273 participants (60% with insufficient and 40% with sufficient cumulative average MVLTPA), whose baseline characteristics by MVLTPA category are shown in **Table 1**. The WHtR analyses included 4,940 participants, representing approximately 94% of the BMI analytic sample, and showed a very similar descriptive profile (**Supplementary Table 2**). Across analyses, participants with higher MVLTPA levels consistently exhibited a more favourable sociodemographic and cardiometabolic profile. Participants contributed a median of 4 MVLTPA measurements to the cumulative mean.

**Table 1.**
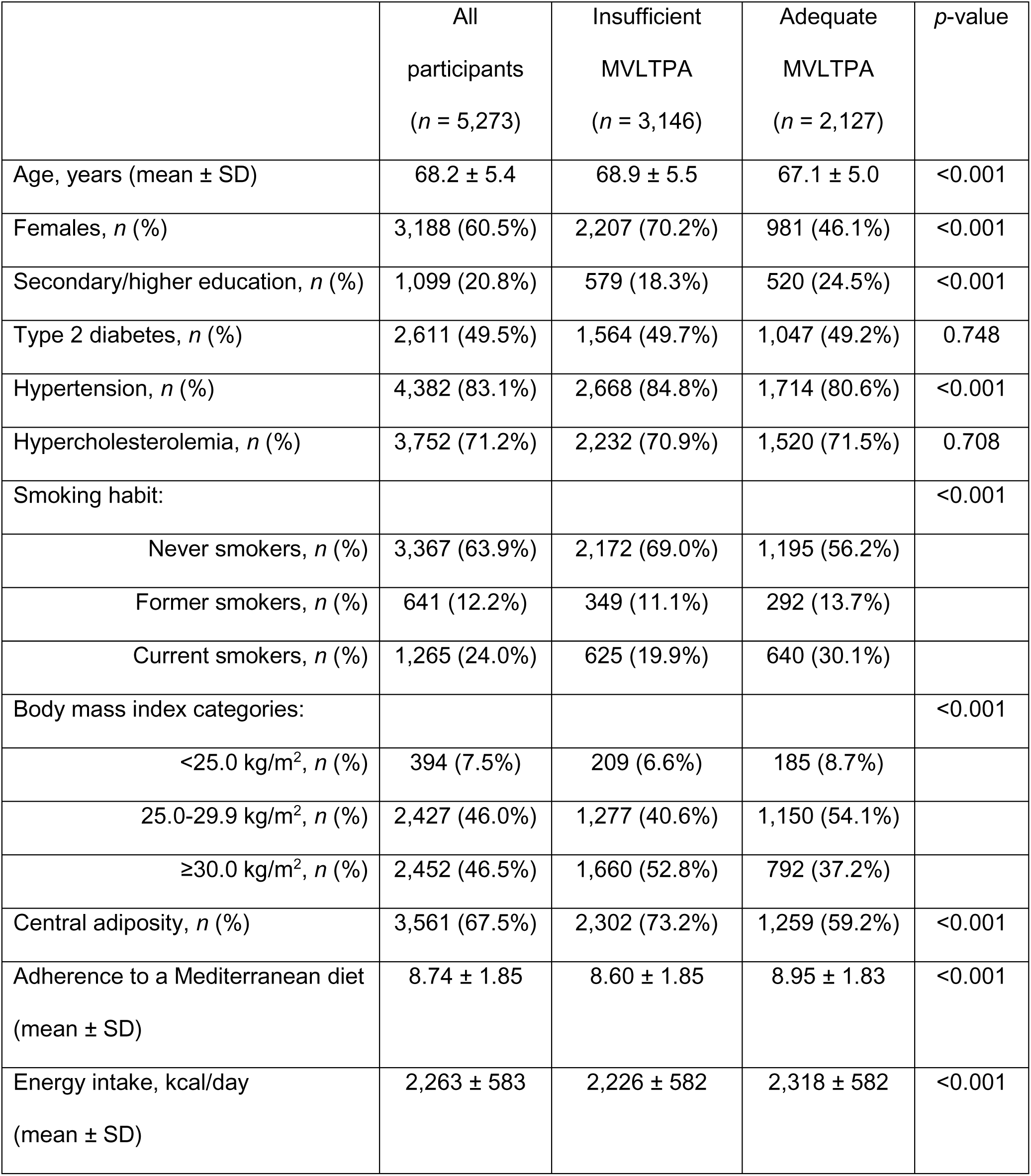
Characteristics of participants in the BMI analyses in the PREDIMED baseline visit.

|  | All<br>participants<br>(*n* = 5,273) | Insufficient<br>MVLTPA<br>(*n* = 3,146) | Adequate<br>MVLTPA<br>(*n* = 2,127) | *p*-value |
| --- | --- | --- | --- | --- |
| Age, years (mean ± SD) | 68.2 ± 5.4 | 68.9 ± 5.5 | 67.1 ± 5.0 | <0.001 |
| Females, *n* (%) | 3,188 (60.5%) | 2,207 (70.2%) | 981 (46.1%) | <0.001 |
| Secondary/higher education, *n* (%) | 1,099 (20.8%) | 579 (18.3%) | 520 (24.5%) | <0.001 |
| Type 2 diabetes, *n* (%) | 2,611 (49.5%) | 1,564 (49.7%) | 1,047 (49.2%) | 0.748 |
| Hypertension, *n* (%) | 4,382 (83.1%) | 2,668 (84.8%) | 1,714 (80.6%) | <0.001 |
| Hypercholesterolemia, *n* (%) | 3,752 (71.2%) | 2,232 (70.9%) | 1,520 (71.5%) | 0.708 |
| Smoking habit: |  |  |  | <0.001 |
| Never smokers, *n* (%) | 3,367 (63.9%) | 2,172 (69.0%) | 1,195 (56.2%) |  |
| Former smokers, *n* (%) | 641 (12.2%) | 349 (11.1%) | 292 (13.7%) |  |
| Current smokers, *n* (%) | 1,265 (24.0%) | 625 (19.9%) | 640 (30.1%) |  |
| Body mass index categories: |  |  |  | <0.001 |
| <25.0 kg/m<sup>2</sup>, *n* (%) | 394 (7.5%) | 209 (6.6%) | 185 (8.7%) |  |
| 25.0-29.9 kg/m<sup>2</sup>, *n* (%) | 2,427 (46.0%) | 1,277 (40.6%) | 1,150 (54.1%) |  |
| ≥30.0 kg/m<sup>2</sup>, *n* (%) | 2,452 (46.5%) | 1,660 (52.8%) | 792 (37.2%) |  |
| Central adiposity, *n* (%) | 3,561 (67.5%) | 2,302 (73.2%) | 1,259 (59.2%) | <0.001 |
| Adherence to a Mediterranean diet<br>(mean ± SD) | 8.74 ± 1.85 | 8.60 ± 1.85 | 8.95 ± 1.83 | <0.001 |
| Energy intake, kcal/day<br>(mean ± SD) | 2,263 ± 583 | 2,226 ± 582 | 2,318 ± 582 | <0.001 |

### Trajectories of BMI and WHtR

BMI tended to decline over time in all participants, but individuals with adequate MVLTPA (compared with those with insufficient MVLTPA) consistently exhibited lower predicted BMI values, with differences between ages 60-80 ranging from-1.27 to-1.48 kg/m² in the full study population (**Figure 2A**). Trajectories were significantly different between women and men (*p*-value for interaction <0.001). Differences were larger in women (ranging from-1.36 to-1.63 kg/m^2^, **Figure 2B**) than in men (ranging from-0.75 to-0.85 kg/m^2^, **Figure 2C**), and smaller among individuals with baseline obesity (ranging from-0.77 to-1.10 kg/m^2^, **Figure 2D**). Exact between-group differences are shown in **Supplementary Figure 1**; exact predicted values represented in the trajectory curves are shown in **Supplementary Table 3**. Individuals with adequate MVLTPA also showed a slower BMI decline from ages 70 to 80 years in the whole study population (insufficient MVLTPA:-0.090 kg/m^2^ per year, 95% CI-0.10 to - 0.079; adequate MVLTPA:-0.073 kg/m^2^ per year, 95% CI-0.081 to-0.064), as well as in women (insufficient:-0.099 kg/m^2^ per year, 95% CI-0.11 to-0.087; adequate:-0.073 kg/m^2^ per year, 95% CI-0.086 to-0.061), and individuals with baseline obesity (insufficient:-0.12 kg/m^2^ per year, 95% CI-0.13 to-0.11; adequate:-0.088 kg/m^2^ per year, 95% CI-0.098 to-0.078), but not in men (insufficient:-0.069 kg/m^2^ per year, 95% CI-0.076 to-0.062; adequate:-0.071 kg/m^2^ per year, 95% CI-0.077 to-0.065).

**Figure 2.**
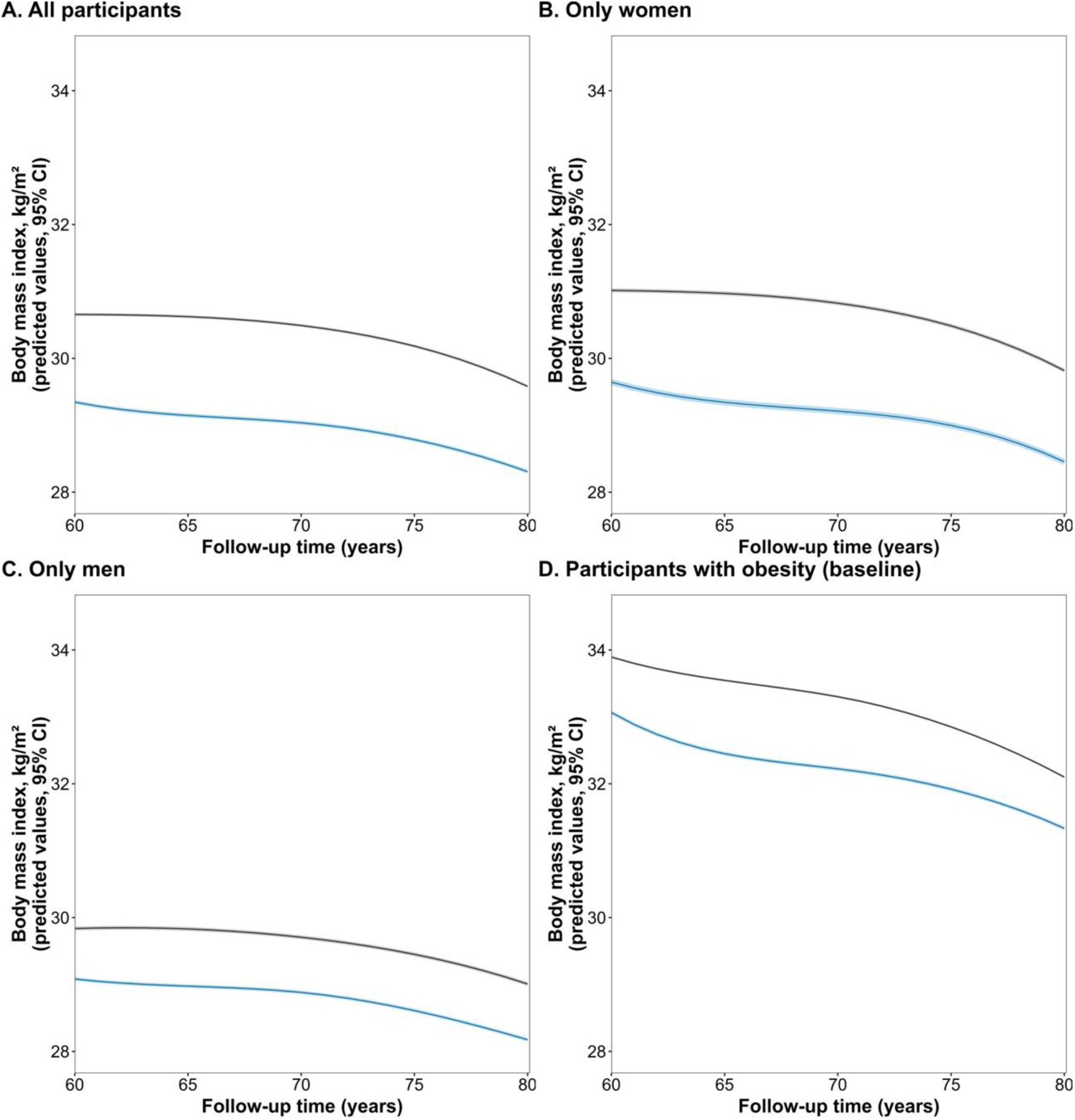
Age-related trajectories of BMI according to cumulative average MVLTPA category. Panels show trajectories (with 95% CI) of predicted mean BMI in all participants (A), women (B), men (C), and participants with obesity at baseline (D). Blue represents participants with adequate MVLTPA (≥100 MET·min/day); grey represents participants with insufficient MVLTPA (<100 MET·min/day).

WHtR increased over time in all participants, except among individuals with central adiposity at baseline. Nevertheless, as observed for BMI, individuals with adequate MVLTPA levels had lower WHtR values between ages 60-80 than those with insufficient MVLTPA, with differences ranging from **-**0.021 to-0.026 in the full study population **(Figure 3A)**. Trajectories were significantly different between women and men (*p*-value for interaction <0.001). Differences were larger in women (ranging from - 0.021 to-0.029, **Figure 3B**) than in men (ranging from-0.014 to-0.019, **Figure 3C**), and smaller among individuals with baseline central adiposity (ranging from-0.013 to - 0.020, **Figure 3D)**. Exact between-group differences are available in **Supplementary Figure 2**; exact predicted values represented in the trajectory curves are shown in **Supplementary Table 4**.

**Figure 3.**
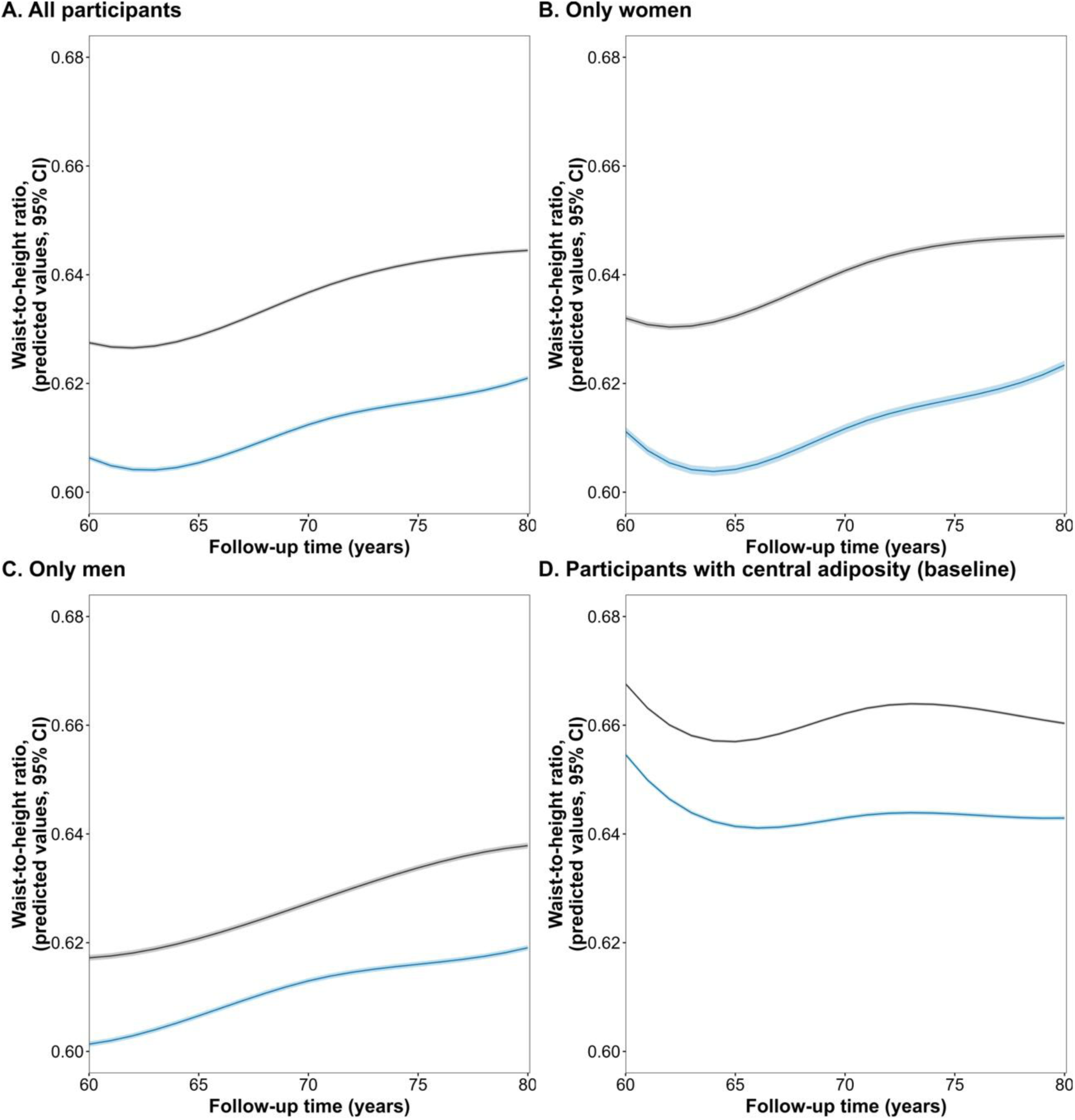
Age-related trajectories of waist-to-height ratio according to cumulative average MVLTPA category. Panels show trajectories (with 95% CI) of waist-to-height ratio in all participants (A), women (B), men (C), and participants with central adiposity at baseline (D). Blue represents participants with adequate MVLTPA (≥100 MET·min/day); grey represents participants with insufficient MVLTPA (<100 MET·min/day).

### Onset of obesity and central adiposity

Among participants without obesity at the baseline PREDIMED visit, first occurrence of overall obesity was observed in 15.1% of participants between 60 and 80 years of age. Adequate MVLTPA was not significantly associated with incident obesity in the total study population (HR 0.91, 95% CI 0.74 to 1.13). However, there was suggestive evidence of an interaction by sex (*p*-value for interaction = 0.084), with divergent point estimates (women: HR 0.76, 95% CI 0.57 to 1.00; men: HR 1.14, 95% CI 0.82 to 1.58; **Figure 4B-C**). Dose-response analyses did not support a significant linear or non-linear association between MVLTPA and incident obesity, with no evidence of differences by sex (**Figure 4D-E**).

**Figure 4.**
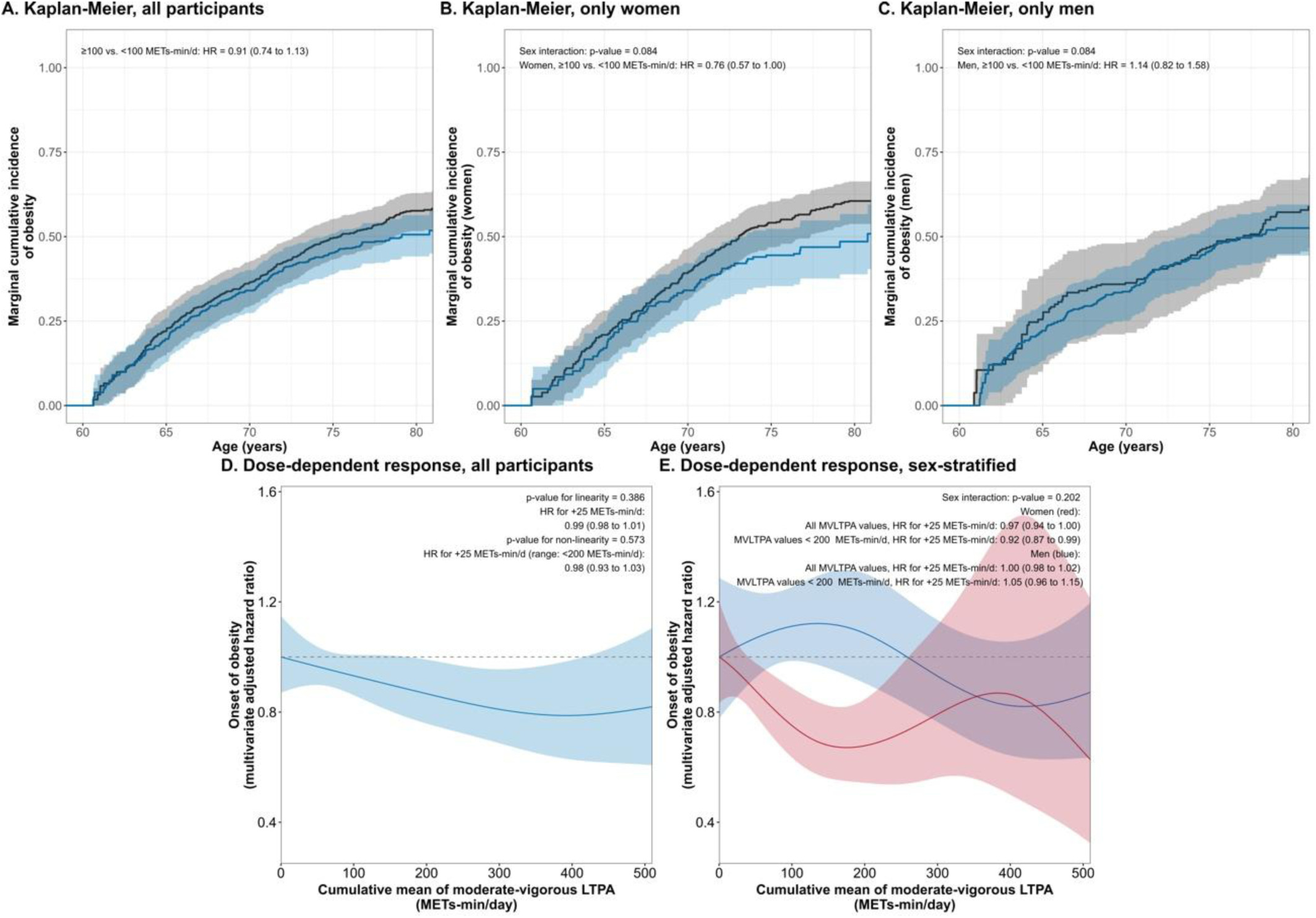
Association between cumulative average MVLTPA and incident BMI-defined obesity between 60 and 80 years of age. Panels show Kaplan-Meier curves in all participants (A), women (B), and men (C); the dose-response association in the overall population (D), and sex-specific dose-response associations (E). In panels A-C, blue represents participants with adequate MVLTPA (≥100 MET·min/day) and grey represents participants with insufficient MVLTPA (<100 MET·min/day). In panel E, red represents women and blue represents men.

Among participants without central adiposity at the baseline PREDIMED visit, first occurrence of central adiposity was observed in 42.4% of participants between 60 and 80 years of age. Adequate MVLTPA was associated with a 34% lower risk of incident central adiposity (HR 0.66, 95% CI 0.56 to 0.77; **Figure 5A**). Similar associations were observed in women and men, with no significant sex interaction (p-value = 0.643; **Figures 5B-5C**). Dose-response analyses showed a significant inverse, non-linear association (*p*-value for non-linearity = 0.021), with progressively lower risk of central adiposity at increasing MVLTPA levels and approximately 40% lower risk at 400 MET·min/day, above which the association plateaued (**Figure 5D**). This dose-response pattern did not differ significantly between women and men (*p*-value for interaction = 0.883; **Figure 5E**).

**Figure 5.**
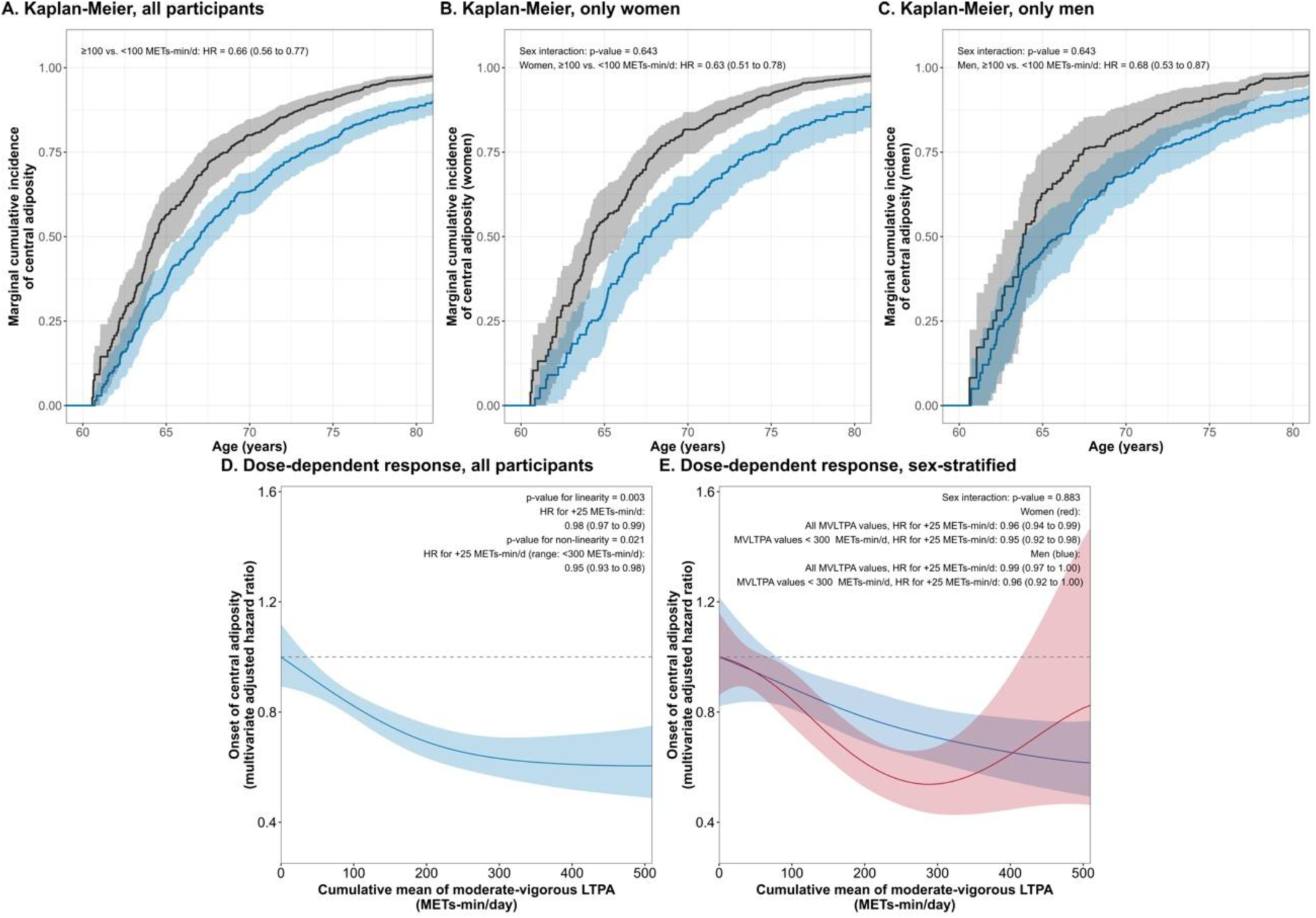
Association between cumulative average MVLTPA and incident central adiposity between 60 and 80 years of age. Panels show Kaplan-Meier curves in all participants (A), women (B), and men (C); the dose-response association in the overall population (D), and sex-specific dose-response associations (E). In panels A-C, blue represents participants with adequate MVLTPA (≥100 MET·min/day) and grey represents participants with insufficient MVLTPA (<100 MET·min/day). In panel E, red represents women and blue represents men.

## DISCUSSION

In this prospective cohort of older adults at high cardiovascular risk, participants with adequate MVLTPA (≥100 MET·min/day) exhibited lower predicted BMI and WHtR values between 60 and 80 years of age, and a slower BMI decline between 70 and 80 years, compared with those with insufficient MVLTPA. Differences in BMI and WHtR predicted mean trajectories were more pronounced in women than in men. Adequate MVLTPA was associated with a lower risk of incident central adiposity. For obesity, there was suggestive evidence of an interaction by sex, with an inverse association in women but not in men. Dose-response analyses showed a significant, non-linear inverse association between MVLTPA and central adiposity risk, with approximately 40% lower risk at 400 MET·min/day and a plateau at higher levels, but no significant dose-response pattern for obesity. Our study complements shorter-term evidence from randomized trials (5), helps address key gaps in the literature on MVLTPA and adiposity in older adults, and shows that these associations differ by adiposity marker, age, and sex.

The BMI findings should be interpreted in the context of late-life body composition changes and the limitations of BMI, which does not differentiate between fat and lean mass and may inadequately reflect age-related changes in body composition, including sarcopenia and fat redistribution (9–11). In our study, participants with adequate MVLTPA had lower BMI values but also showed a slower BMI decline between 70 and 80 years of age. Lower BMI may partly reflect a more favourable anthropometric profile established earlier in adulthood, whereas the slower decline in body mass may be compatible with less pronounced age-related loss of body mass, although BMI alone does not allow the underlying changes in fat and lean mass to be determined (24). The more limited incidence of BMI-defined obesity in this group (∼15%) could partly explain the lack of robust associations between MVLTPA and incident obesity in our data. The lower BMI and slower BMI decline observed with adequate MVLTPA are broadly consistent with previous evidence of an association between physical activity and more favourable weight trajectories in older adults (25, 26).

In contrast, WHtR has been proposed as a more sensitive marker of central adiposity that may better capture cardiometabolic risk than BMI (27, 28). Most previous longitudinal studies of central adiposity in older adults have relied on waist circumference (29, 30), which uses sex-specific cut-offs to define central obesity. WHtR, by accounting for height, may be less influenced by age-related changes in body composition (31) and provides a single cut-off for central adiposity in both women and men (19). This may partly explain why longitudinal evidence on WHtR trajectories, particularly age-and sex-specific trajectories, remains limited. In our cohort, WHtR increased over time in all participants, but those with adequate MVLTPA had lower WHtR values and a substantially lower risk of incident central adiposity (∼34% less), suggesting that MVLTPA is more clearly associated with waist-related adiposity than with BMI-defined obesity. This is clinically relevant because older adults may accumulate central fat even when BMI is stable or declining (27, 28, 32). Because BMI cannot distinguish fat mass, lean mass, and body water, the slower BMI decline observed in participants with adequate MVLTPA cannot, on its own, be attributed to preserved muscle mass or reduced visceral fat. Prior studies using direct body-composition measures have shown that physical activity in older adults is prospectively associated with preserved lean muscle (33) and reduced visceral adipose tissue (generally linked to central adiposity) despite modest changes in body weight (34). These findings provide a plausible context for our observed BMI and WHtR patterns, although detailed body-composition measures were not available in our study. Finally, our dose-response analyses suggested that higher MVLTPA levels were associated with progressively lower central adiposity risk up to approximately 400 MET·min/day, where the risk was approximately 40% lower and the association plateaued thereafter. This level extends beyond the minimum recommended amount of MVLTPA (6, 7). These findings support meeting current physical activity recommendations for cardiovascular prevention in older adults while suggesting that MVLTPA levels beyond the recommendations (up to twice the current recommended levels) may provide additional benefits for central adiposity.

Sex differences in the associations between MVLTPA and adiposity are suggested in our data. For BMI and WHtR trajectories, women showed more pronounced associations and a slower late-life BMI decline, and a suggestive association with lower risk of overall obesity that was not observed in men. From a prevention perspective, this is relevant because adherence to physical activity recommendations is generally lower among older women than men (35, 36), supporting the need for sex-sensitive strategies to promote MVLTPA in older adults. These sex differences may partly reflect known differences in fat distribution between women and men, including a shift toward more central fat deposition after menopause (37), though the mechanisms remain incompletely understood.

This study has several limitations. First, as our analyses are observational, causal conclusions regarding the effect of MVLTPA on adiposity cannot be drawn, and the observed associations may be partly explained by residual confounding despite adjustment for multiple covariates. Reverse causation cannot be excluded: lower MVLTPA levels in older adults may partly reflect underlying frailty, mobility limitations, or sarcopenia, which themselves influence adiposity. Our findings should be interpreted as complementary to randomized trial evidence, extending it by suggesting that favorable associations between MVLTPA and adiposity may persist over longer follow-up and across older age. Future interventional studies or triangulation with complementary epidemiological approaches (12, 38) are warranted to confirm our results. Second, physical activity was assessed using a self-reported questionnaire, which may be subject to recall bias and measurement error. However, the use of cumulative averages of repeated measurements likely reduced random variability and better captured long-term exposure. Third, the longitudinal database was reconstructed from a discontinuously observed panel rather than continuous follow-up of all individuals across the entire age range, so trajectory estimates at different ages may be informed by different subsets of participants. We minimized this through: 1) mixed-effects spline models using all available repeated measurements; 2) restricting trajectory analyses to participants with at least two measurements; 3) inverse probability weighting in Kaplan-Meier analyses to address missingness; and 4) age-as-time-scale Cox models treating loss to follow-up as right censoring. In addition, analyses in subgroups (women, men, and participants with excess adiposity at baseline) yielded results consistent with the main findings. Nevertheless, these approaches assume missing data are missing at random and loss to follow-up is non-informative after covariate adjustment; violations could bias the estimates, and prospective studies with complete long-term follow-up are still needed. Finally, the study population consisted of older adults at high cardiovascular risk from a Mediterranean setting, which may limit the generalizability of the findings to younger populations, lower-risk individuals, or non-Mediterranean populations.

## CONCLUSION

In older adults at high cardiovascular risk, higher cumulative MVLTPA was associated with more favourable adiposity profiles between 60 and 80 years of age, particularly for WHtR-defined central adiposity. Adequate MVLTPA was linked to lower BMI and WHtR values, slower late-life BMI decline, and a lower risk of incident central adiposity, with dose-response analyses suggesting the greatest benefit up to 400 MET·min/day. Our study complements shorter-term randomized trial evidence and extends its conclusions to longer age ranges and more detailed population subgroups. These findings support promoting MVLTPA for cardiovascular and chronic disease prevention in older adults, particularly among inactive individuals, and support the use of waist-based indicators to monitor activity-related adiposity benefits. Future studies should confirm these findings in independent cohorts and intervention settings and evaluate whether similar beneficial associations with MVLTPA are also observed for other cardiometabolic risk factors.

## ACKNOWLEDGMENTS

We gratefully acknowledge all PREDIMED participants, investigators, and study personnel involved in participant recruitment, follow-up, data collection, and study coordination. CIBER de Enfermedades Cardiovasculares (CIBERCV) and CIBER de Fisiopatología de la Obesidad y Nutrición (CIBEROBN) are initiatives of Instituto de Salud Carlos III (Madrid, Spain) and are financed by the European Regional Development Fund.

## AUTHOR CONTRIBUTIONS

Conceptualization: Á.H. Methodology: E.F., Á.H.. Formal analysis: E.F., Á.H. Investigation: A.G., M.Á.M.-G., J.S.-S., R.E., E.R., E.G.-G., M.Fiol, J.L., J.T., L.S.-M., X.P., M.Fitó. Data curation: Á.H. Writing-original draft: E.F. Writing-review and editing: M.H.H., A.G., A.C.-V., M.Á.M.-G., J.S.-S., R.E., E.R., E.G.-G., M.Fiol, J.L., J.T., L.S.- M., X.P., Z.V.-R., F.M.M.-L., M.Fitó, Á.H. Funding acquisition: Á.H. Supervision: M.H.H., Á.H. All authors read and approved the final manuscript.

## DATA AVAILABILITY

Because national data-protection legislation and ethical restrictions prohibit public dissemination of the individual-level PREDIMED dataset, and because participants did not provide explicit written consent for open data sharing, the study dataset cannot be deposited in a public repository. Access to data for bona fide collaborative research may be facilitated for qualified investigators upon reasonable request and approval by the PREDIMED Steering Committee. Requests should be addressed to. The code used for constructing the database, managing the data, and performing the statistical analyses is openly available at: https://github.com/alvarohernaez/MVLTPA_cardiometab_PREDIMED.

