## Supplemental Materials for "Moderate-to-vigorous physical activity, age-dependent adiposity, and obesity risk in older adults at high cardiovascular risk"

SUPPLEMENTARY MATERIALS

SUPPLEMENTARY TABLES

**Supplementary Table 1.** Number of BMI and WHtR observations available at each chronological age between 60 and 80 years.

| Age | BMI measures | WHtR measures |
|---|---|---|
| 60 | 339 | 322 |
| 61 | 593 | 565 |
| 62 | 835 | 776 |
| 63 | 1,067 | 996 |
| 64 | 1,206 | 1,133 |
| 65 | 1,352 | 1,250 |
| 66 | 1,400 | 1,298 |
| 67 | 1,451 | 1,346 |
| 68 | 1,406 | 1,291 |
| 69 | 1,401 | 1,278 |
| 70 | 1,368 | 1,243 |
| 71 | 1,357 | 1,254 |
| 72 | 1,419 | 1,297 |
| 73 | 1,341 | 1,231 |
| 74 | 1,217 | 1,112 |
| 75 | 1,143 | 1,059 |
| 76 | 1,061 | 984 |
| 77 | 904 | 825 |
| 78 | 817 | 731 |
| 79 | 662 | 600 |
| 80 | 517 | 455 |

**Supplementary Table 2**. Characteristics of participants in the waist-to-height ratio analyses in the PREDIMED baseline visit.

| | All participants | Insufficient MVLTPA | Adequate MVLTPA |
|---|---|---|---|
| Age, years (mean ± SD) | 68.1 ± 5.39 | 68.8 ± 5.54 | 67.1 ± 4.98 |
| Females, *n* (%) | 2,965 (60.0%) | 2,040 (70.2%) | 925 (45.5%) |
| Secondary/higher education, *n* (%) | 1,033 (20.9%) | 534 (18.4%) | 497 (24.4%) |
| Type-II diabetes, *n* (%) | 2,417 (48.9%) | 1,422 (48.9%) | 995 (48.9%) |
| Hypertension, *n* (%) | 4,103 (83.1%) | 2,466 (84.8%) | 1,637 (80.5%) |
| Hypercholesterolemia, *n* (%) | 3,513 (71.1%) | 2,061 (70.9%) | 1,452 (71.4%) |
| Hypertriglyceridemia, *n* (%) | 1,416 (28,%) | 882 (30,3%) | 534 (26.3%) |
| Smoking habit: | | | |
| Never smokers, *n* (%) | 3,149 (63.7%) | 2,008 (69.1%) | 1,141 (56.1%) |
| Former smokers, *n* (%) | 600 (12.1%) | 326 (11.2%) | 274 (13.5%) |
| Current smokers, *n* (%) | 1,191 (24.1%) | 573 (19.7%) | 618 (30.4%) |
| Body mass index categories: | | | |
| <25.0 kg/m$^2$, *n* (%) | 380 (7.69%) | 199 (6.85%) | 181 (8.90%) |
| 25.0-29.9 kg/m$^2$, *n* (%) | 2,293 (46.4%) | 1,192 (41.0%) | 1,101 (54.2%) |
| ≥30.0 kg/m$^2$, *n* (%) | 2,267 (45.9%) | 1,516 (52.1%) | 751 (36.9%) |
| Central adiposity, *n* (%) | 3,304 (66.9%) | 2,112 (72.7%) | 1,192 (58.6%) |
| Adherence to a Mediterranean diet (mean ± SD) | 8.75 ± 1.86 | 8.59 ± 1.85 | 8.97 ± 1.85 |
| Energy intake, kcal/day (mean ± SD) | 2,269 ± 583 | 2,232 ± 582 | 2,322 ± 580 |

**Supplementary Table 3.** Exact predicted mean values and their 95% CI presented in the BMI trajectory curves in all ages between 60 and 80.

| Age | 60 | 61 | 62 | 63 | 64 | 65 | 66 | 67 | 68 | 69 |
|---|---|---|---|---|---|---|---|---|---|---|
| Insufficient MVLTPA, all participants | 30.656 (30.633 to 30.679) | 30.654 (30.631 to 30.677) | 30.651 (30.628 to 30.674) | 30.645 (30.622 to 30.668) | 30.636 (30.613 to 30.659) | 30.623 (30.601 to 30.646) | 30.607 (30.584 to 30.630) | 30.586 (30.563 to 30.609) | 30.561 (30.538 to 30.583) | 30.529 (30.506 to 30.552) |
| Sufficient MVLTPA, all participants | 29.346 (29.317 to 29.376) | 29.287 (29.257 to 29.316) | 29.239 (29.209 to 29.268) | 29.201 (29.171 to 29.231) | 29.171 (29.141 to 29.200) | 29.146 (29.117 to 29.176) | 29.125 (29.096 to 29.155) | 29.106 (29.076 to 29.136) | 29.086 (29.057 to 29.116) | 29.064 (29.034 to 29.094) |
| Insufficient MVLTPA, only women | 31.016 (30.987 to 31.046) | 31.011 (30.981 to 31.041) | 31.005 (30.975 to 31.034) | 30.996 (30.966 to 31.026) | 30.985 (30.955 to 31.015) | 30.971 (30.941 to 31.001) | 30.952 (30.923 to 30.982) | 30.929 (30.900 to 30.959) | 30.901 (30.871 to 30.931) | 30.866 (30.837 to 30.896) |
| Sufficient MVLTPA, only women | 29.645 (29.598 to 29.692) | 29.559 (29.512 to 29.606) | 29.488 (29.441 to 29.536) | 29.430 (29.383 to 29.477) | 29.383 (29.335 to 29.430) | 29.344 (29.297 to 29.391) | 29.312 (29.265 to 29.359) | 29.285 (29.237 to 29.332) | 29.260 (29.213 to 29.308) | 29.237 (29.189 to 29.284) |
| Insufficient MVLTPA, only men | 29.837 (29.808 to 29.866) | 29.845 (29.816 to 29.873) | 29.848 (29.819 to 29.877) | 29.847 (29.818 to 29.876) | 29.841 (29.812 to 29.870) | 29.830 (29.802 to 29.859) | 29.815 (29.786 to 29.844) | 29.795 (29.766 to 29.824) | 29.771 (29.742 to 29.800) | 29.741 (29.712 to 29.770) |
| Sufficient MVLTPA, only men | 29.082 (29.054 to 29.109) | 29.048 (29.021 to 29.075) | 29.023 (28.996 to 29.050) | 29.004 (28.976 to 29.031) | 28.988 (28.961 to 29.016) | 28.976 (28.948 to 29.003) | 28.963 (28.936 to 28.990) | 28.950 (28.922 to 28.977) | 28.933 (28.906 to 28.960) | 28.911 (28.884 to 28.938) |
| Insufficient MVLTPA, participants with obesity at baseline | 33.894 (33.875 to 33.912) | 33.799 (33.780 to 33.817) | 33.719 (33.701 to 33.738) | 33.653 (33.634 to 33.671) | 33.595 (33.577 to 33.614) | 33.545 (33.527 to 33.564) | 33.499 (33.481 to 33.518) | 33.454 (33.436 to 33.473) | 33.408 (33.390 to 33.427) | 33.358 (33.340 to 33.377) |
| Sufficient MVLTPA, participants with obesity at baseline | 33.065 (33.036 to 33.094) | 32.886 (32.857 to 32.915) | 32.739 (32.710 to 32.768) | 32.620 (32.591 to 32.649) | 32.525 (32.496 to 32.554) | 32.449 (32.421 to 32.478) | 32.389 (32.361 to 32.418) | 32.341 (32.312 to 32.370) | 32.300 (32.271 to 32.329) | 32.262 (32.233 to 32.291) |

| Age | 70 | 71 | 72 | 73 | 74 | 75 | 76 | 77 | 78 | 79 | 80 |
|---|---|---|---|---|---|---|---|---|---|---|---|
| Insufficient MVLTPA, all participants | 30.491 (30.468 to 30.514) | 30.447 (30.424 to 30.470) | 30.395 (30.372 to 30.418) | 30.335 (30.312 to 30.357) | 30.264 (30.241 to 30.287) | 30.184 (30.161 to 30.206) | 30.091 (30.068 to 30.114) | 29.985 (29.963 to 30.008) | 29.866 (29.843 to 29.889) | 29.732 (29.709 to 29.755) | 29.581 (29.558 to 29.604) |
| Sufficient MVLTPA, all participants | 29.037 (29.007 to 29.067) | 29.004 (28.974 to 29.033) | 28.962 (28.932 to 28.992) | 28.912 (28.882 to 28.942) | 28.854 (28.824 to 28.883) | 28.786 (28.757 to 28.816) | 28.710 (28.680 to 28.740) | 28.624 (28.594 to 28.654) | 28.529 (28.499 to 28.558) | 28.423 (28.394 to 28.453) | 28.308 (28.278 to 28.337) |
| Insufficient MVLTPA, only women | 30.825 (30.795 to 30.855) | 30.776 (30.746 to 30.806) | 30.719 (30.689 to 30.749) | 30.652 (30.623 to 30.682) | 30.575 (30.545 to 30.605) | 30.486 (30.456 to 30.516) | 30.383 (30.354 to 30.413) | 30.267 (30.237 to 30.296) | 30.134 (30.105 to 30.164) | 29.985 (29.955 to 30.015) | 29.818 (29.788 to 29.848) |
| Sufficient MVLTPA, only women | 29.212 (29.164 to 29.259) | 29.183 (29.136 to 29.231) | 29.150 (29.103 to 29.197) | 29.109 (29.062 to 29.156) | 29.059 (29.012 to 29.107) | 28.998 (28.951 to 29.045) | 28.924 (28.876 to 28.971) | 28.834 (28.787 to 28.882) | 28.728 (28.680 to 28.775) | 28.602 (28.555 to 28.649) | 28.455 (28.408 to 28.502) |
| Insufficient MVLTPA, only men | 29.707 (29.678 to 29.736) | 29.667 (29.638 to 29.696) | 29.623 (29.594 to 29.651) | 29.572 (29.543 to 29.601) | 29.515 (29.486 to 29.544) | 29.451 (29.422 to 29.480) | 29.380 (29.351 to 29.409) | 29.300 (29.271 to 29.329) | 29.212 (29.183 to 29.241) | 29.115 (29.086 to 29.144) | 29.008 (28.979 to 29.037) |
| Sufficient MVLTPA, only men | 28.882 (28.855 to 28.909) | 28.845 (28.817 to 28.872) | 28.798 (28.771 to 28.825) | 28.743 (28.716 to 28.770) | 28.680 (28.653 to 28.707) | 28.610 (28.583 to 28.637) | 28.534 (28.506 to 28.561) | 28.451 (28.424 to 28.478) | 28.364 (28.336 to 28.391) | 28.271 (28.244 to 28.298) | 28.175 (28.148 to 28.202) |
| Insufficient MVLTPA, participants with obesity at baseline | 33.301 (33.282 to 33.319) | 33.234 (33.215 to 33.252) | 33.155 (33.137 to 33.174) | 33.066 (33.047 to 33.084) | 32.964 (32.945 to 32.982) | 32.850 (32.832 to 32.869) | 32.725 (32.706 to 32.744) | 32.587 (32.569 to 32.606) | 32.437 (32.419 to 32.456) | 32.275 (32.257 to 32.294) | 32.100 (32.082 to 32.119) |
| Sufficient MVLTPA, participants with obesity at baseline | 32.223 (32.194 to 32.252) | 32.179 (32.150 to 32.208) | 32.128 (32.099 to 32.157) | 32.068 (32.039 to 32.097) | 31.999 (31.970 to 32.028) | 31.919 (31.890 to 31.948) | 31.828 (31.799 to 31.857) | 31.725 (31.696 to 31.754) | 31.609 (31.580 to 31.638) | 31.479 (31.450 to 31.508) | 31.334 (31.305 to 31.363) |

**Supplementary Table 4.** Exact predicted mean values and their 95% CI presented in the waist-to-height trajectory curves in all ages between 60 and 80.

| Age | 60 | 61 | 62 | 63 | 64 | 65 | 66 | 67 | 68 | 69 |
|---|---|---|---|---|---|---|---|---|---|---|
| Insufficient MVLTPA, all participants | 0.627 (0.627 to 0.628) | 0.627 (0.626 to 0.627) | 0.627 (0.626 to 0.627) | 0.627 (0.627 to 0.627) | 0.628 (0.627 to 0.628) | 0.629 (0.628 to 0.629) | 0.630 (0.630 to 0.631) | 0.632 (0.631 to 0.632) | 0.633 (0.633 to 0.634) | 0.635 (0.635 to 0.635) |
| Sufficient MVLTPA, all participants | 0.606 (0.606 to 0.607) | 0.605 (0.604 to 0.605) | 0.604 (0.604 to 0.605) | 0.604 (0.604 to 0.605) | 0.605 (0.604 to 0.605) | 0.605 (0.605 to 0.606) | 0.607 (0.606 to 0.607) | 0.608 (0.608 to 0.608) | 0.610 (0.609 to 0.610) | 0.611 (0.611 to 0.611) |
| Insufficient MVLTPA, only women | 0.632 (0.631 to 0.633) | 0.631 (0.630 to 0.631) | 0.630 (0.630 to 0.631) | 0.631 (0.630 to 0.631) | 0.631 (0.631 to 0.632) | 0.632 (0.632 to 0.633) | 0.634 (0.633 to 0.634) | 0.636 (0.635 to 0.636) | 0.637 (0.637 to 0.638) | 0.639 (0.639 to 0.640) |
| Sufficient MVLTPA, only women | 0.611 (0.610 to 0.612) | 0.608 (0.607 to 0.609) | 0.605 (0.605 to 0.606) | 0.604 (0.603 to 0.605) | 0.604 (0.603 to 0.605) | 0.604 (0.603 to 0.605) | 0.605 (0.604 to 0.606) | 0.607 (0.606 to 0.607) | 0.608 (0.607 to 0.609) | 0.610 (0.609 to 0.611) |
| Insufficient MVLTPA, only men | 0.617 (0.617 to 0.618) | 0.618 (0.617 to 0.618) | 0.618 (0.618 to 0.619) | 0.619 (0.618 to 0.619) | 0.620 (0.619 to 0.620) | 0.621 (0.620 to 0.621) | 0.622 (0.621 to 0.622) | 0.623 (0.623 to 0.624) | 0.624 (0.624 to 0.625) | 0.626 (0.625 to 0.626) |
| Sufficient MVLTPA, only men | 0.601 (0.601 to 0.602) | 0.602 (0.601 to 0.602) | 0.603 (0.602 to 0.603) | 0.604 (0.603 to 0.604) | 0.605 (0.605 to 0.606) | 0.607 (0.606 to 0.607) | 0.608 (0.607 to 0.608) | 0.609 (0.609 to 0.610) | 0.611 (0.610 to 0.611) | 0.612 (0.611 to 0.612) |
| Insufficient MVLTPA, participants with c. adiposity at baseline | 0.668 (0.667 to 0.668) | 0.663 (0.663 to 0.663) | 0.660 (0.660 to 0.660) | 0.658 (0.658 to 0.658) | 0.657 (0.657 to 0.657) | 0.657 (0.657 to 0.657) | 0.657 (0.657 to 0.658) | 0.658 (0.658 to 0.659) | 0.660 (0.659 to 0.660) | 0.661 (0.661 to 0.661) |
| Sufficient MVLTPA, participants with c. adiposity at baseline | 0.655 (0.654 to 0.655) | 0.650 (0.650 to 0.650) | 0.646 (0.646 to 0.647) | 0.644 (0.644 to 0.644) | 0.642 (0.642 to 0.643) | 0.641 (0.641 to 0.642) | 0.641 (0.641 to 0.641) | 0.641 (0.641 to 0.642) | 0.642 (0.641 to 0.642) | 0.642 (0.642 to 0.643) |

| Age | 70 | 71 | 72 | 73 | 74 | 75 | 76 | 77 | 78 | 79 | 80 |
|---|---|---|---|---|---|---|---|---|---|---|---|
| Insufficient MVLTPA, all participants | 0.637 (0.636 to 0.637) | 0.638 (0.638 to 0.639) | 0.639 (0.639 to 0.640) | 0.641 (0.640 to 0.641) | 0.642 (0.641 to 0.642) | 0.642 (0.642 to 0.643) | 0.643 (0.643 to 0.643) | 0.643 (0.643 to 0.644) | 0.644 (0.644 to 0.644) | 0.644 (0.644 to 0.645) | 0.644 (0.644 to 0.645) |
| Sufficient MVLTPA, all participants | 0.612 (0.612 to 0.613) | 0.614 (0.613 to 0.614) | 0.615 (0.614 to 0.615) | 0.615 (0.615 to 0.616) | 0.616 (0.616 to 0.616) | 0.617 (0.616 to 0.617) | 0.617 (0.617 to 0.618) | 0.618 (0.617 to 0.618) | 0.619 (0.618 to 0.619) | 0.620 (0.619 to 0.620) | 0.621 (0.621 to 0.621) |
| Insufficient MVLTPA, only women | 0.641 (0.640 to 0.641) | 0.642 (0.642 to 0.643) | 0.643 (0.643 to 0.644) | 0.644 (0.644 to 0.645) | 0.645 (0.645 to 0.646) | 0.646 (0.645 to 0.646) | 0.646 (0.646 to 0.647) | 0.647 (0.646 to 0.647) | 0.647 (0.646 to 0.647) | 0.647 (0.646 to 0.647) | 0.647 (0.647 to 0.648) |
| Sufficient MVLTPA, only women | 0.612 (0.611 to 0.613) | 0.613 (0.612 to 0.614) | 0.614 (0.614 to 0.615) | 0.615 (0.615 to 0.616) | 0.616 (0.616 to 0.617) | 0.617 (0.616 to 0.618) | 0.618 (0.617 to 0.619) | 0.619 (0.618 to 0.620) | 0.620 (0.619 to 0.621) | 0.622 (0.621 to 0.622) | 0.623 (0.623 to 0.624) |
| Insufficient MVLTPA, only men | 0.627 (0.627 to 0.628) | 0.629 (0.628 to 0.629) | 0.630 (0.629 to 0.631) | 0.631 (0.631 to 0.632) | 0.633 (0.632 to 0.633) | 0.634 (0.633 to 0.634) | 0.635 (0.634 to 0.635) | 0.636 (0.635 to 0.636) | 0.637 (0.636 to 0.637) | 0.637 (0.637 to 0.638) | 0.638 (0.637 to 0.638) |
| Sufficient MVLTPA, only men | 0.613 (0.612 to 0.613) | 0.614 (0.613 to 0.614) | 0.615 (0.614 to 0.615) | 0.615 (0.615 to 0.616) | 0.616 (0.615 to 0.616) | 0.616 (0.616 to 0.617) | 0.616 (0.616 to 0.617) | 0.617 (0.616 to 0.617) | 0.617 (0.617 to 0.618) | 0.618 (0.618 to 0.619) | 0.619 (0.619 to 0.620) |
| Insufficient MVLTPA, participants with c.adiposity at baseline | 0.662 (0.662 to 0.662) | 0.663 (0.663 to 0.663) | 0.664 (0.663 to 0.664) | 0.664 (0.664 to 0.664) | 0.664 (0.664 to 0.664) | 0.664 (0.663 to 0.664) | 0.663 (0.663 to 0.663) | 0.662 (0.662 to 0.663) | 0.662 (0.661 to 0.662) | 0.661 (0.661 to 0.661) | 0.660 (0.660 to 0.661) |
| Sufficient MVLTPA, participants with c.adiposity at baseline | 0.643 (0.643 to 0.643) | 0.644 (0.643 to 0.644) | 0.644 (0.643 to 0.644) | 0.644 (0.644 to 0.644) | 0.644 (0.643 to 0.644) | 0.644 (0.643 to 0.644) | 0.643 (0.643 to 0.644) | 0.643 (0.643 to 0.644) | 0.643 (0.643 to 0.643) | 0.643 (0.643 to 0.643) | 0.643 (0.643 to 0.643) |

### SUPPLEMENTAL FIGURES

**Supplementary Figure 1.** Inter-group differences in BMI between participants with sufficient and insufficient MVLTPA at ages 60, 65, 70, 75, and 80, presented for the overall sample (black) and stratified by sex (women in maroon, men in green) and baseline obesity status (all individuals with baseline obesity in purple, women in fuchsia, men in orange).

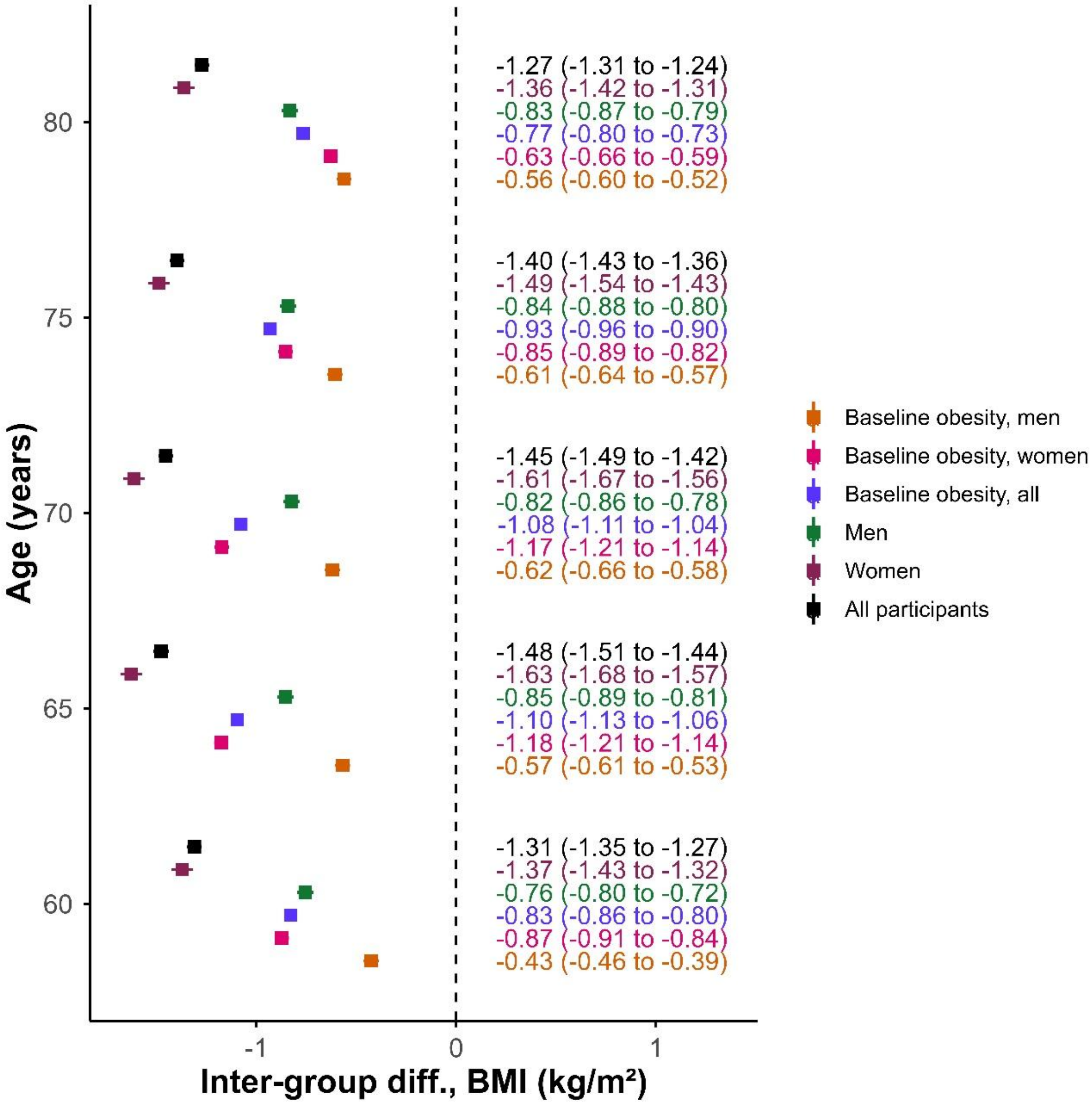

**Supplementary Figure 2.** Inter-group differences in waist-to-height ratio between participants with sufficient and insufficient MVLTPA at ages 60, 65, 70, 75, and 80, presented for the overall sample (black) and stratified by sex (women in maroon, men in green) and baseline obesity status (all individuals with baseline obesity in purple, women in fuchsia, men in orange).

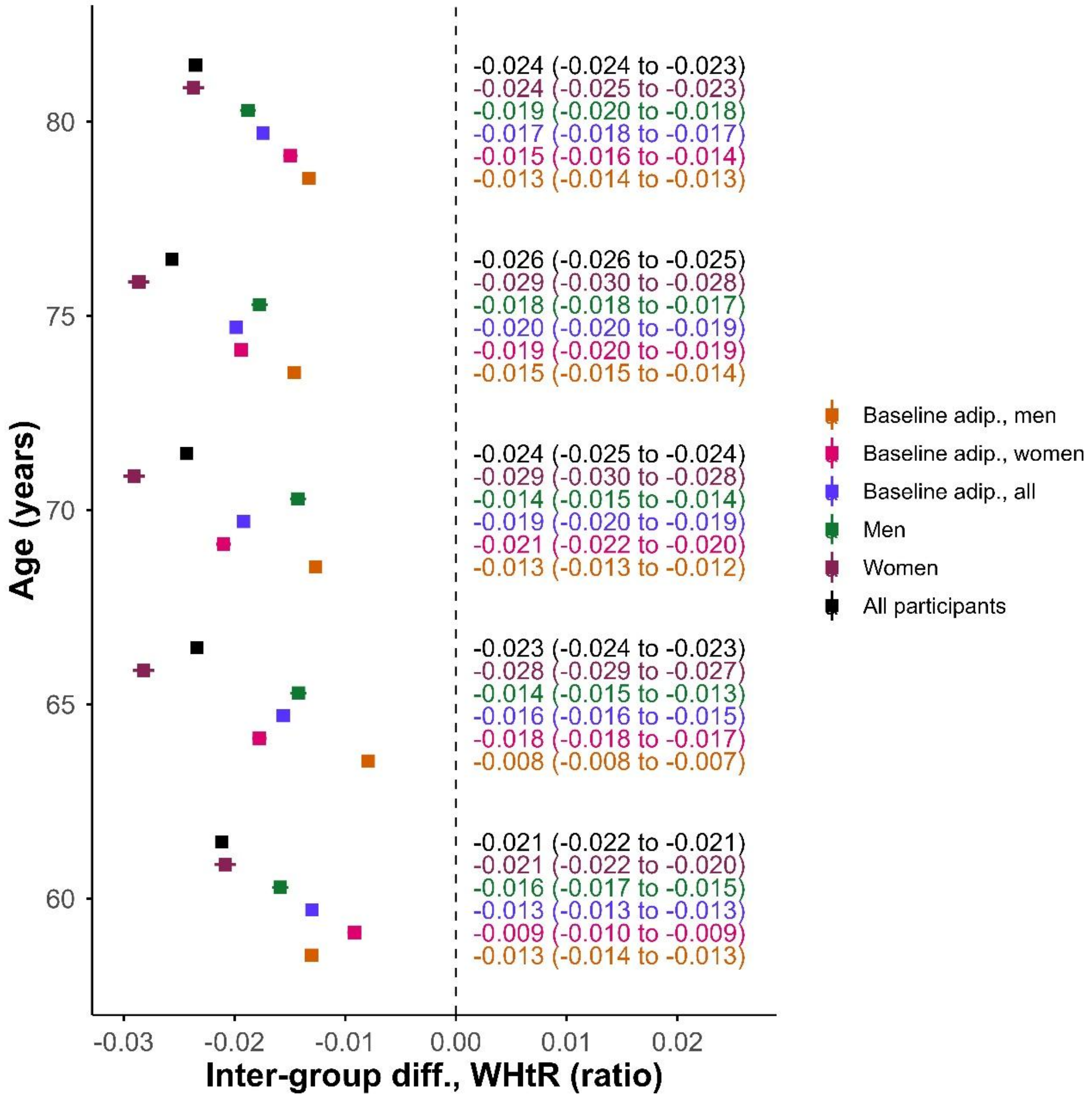
